# Protocol for an eDelphi study to identify consensus on policy metrics which should be included in future assessments of the England Rare Diseases Action Plans

**DOI:** 10.1101/2025.04.26.25326471

**Authors:** Petra A Wark, Leeza Osipenko, Josie Godfrey, Lindsay Birrell, Aimee Walker-Clarke, Andrea Caputo Svensson, Scott Lewis, Jack Lewis, Mark Sheehan, Robin H Lachmann, Paul Gissen, Ala Szczepura

## Abstract

**Introduction:** Governments create policies to address societal needs and then assess their effectiveness through policy metrics. The UK Rare Diseases Framework and its England Rare Diseases Action Plans aim to improve the lives of people with lived experience of rare diseases and their carers. Reaching these goals requires more effective engagement with those least likely to interact with NHS services. This paper outlines a protocol to identify relevant high-level policy metrics while engaging underserved and underrepresented groups.

**Methods and Analysis:** A long list of candidate metrics for the 36 Actions linked to the four Priority Areas of the Framework was compiled from government logic models. The list was discussed in three workshops, and irrelevant, ambiguous, or time-irrelevant metrics were removed. Additional metrics were identified through an evidence review and expert opinion.

In the first round of a two-round eDelphi study, shortlisted metrics will be grouped into the four Priority Areas: Faster Diagnosis, Increased Healthcare Professional Awareness, Better Care Coordination, and Improved Access to Specialist Care, Treatment, and drugs. An eDelphi panel of 50 people living with rare disease or their carers, as well as 50 professional stakeholders, will rate each metric on a 1-9 scale. The sample size allows for 20% attrition. Consensus for inclusion or exclusion will be based on 70% agreement (7-9 for ‘very valuable’, 1-3 for ‘not valuable’), with fewer than 15% rating the metric at the opposite end of the scale.

Metrics will be included or excluded if consensus is reached in both groups; otherwise, they will remain under consideration in Round 2.

In Round 1, eDelphi participants may propose additional metrics, which will be included in Round 2. After two rounds, a final set of policy metrics will be selected. Any discrepancies in Round 2 ratings will be explored through further research.

**Ethics and Dissemination:** The Coventry University Group Research Ethics Committee has approved the study (P183377). The results will be disseminated to people with lived experience of rare disease and professional stakeholders through conferences, events, and newsletters and submitted to a peer-reviewed journal. If adopted, the identified metrics will inform decision-making on rare disease policies in England and beyond.

**Registration Details:** ISRCTN41639707

**ARTICLE SUMMARY:** *Strengths and limitations of this study:*

- The study’s strength lies in its equal representation of individuals with lived experience of rare diseases and professional stakeholders, ensuring contextually relevant and widely accepted policy metrics.
- Efforts are made to include underserved groups, ensuring the meaningful representation of underrepresented sub-populations.
- The findings will primarily apply to the England Rare Diseases Action Plans, limiting their generalisability to other countries or healthcare systems.
- The heterogeneity of the study population may pose challenges in reaching consensus, potentially impacting the robustness of the agreed-policy metrics.
- Despite targeted recruitment efforts, challenges in including underserved groups may result in a less representative sample, affecting the study’s generalisability.

## 1. INTRODUCTION

Governmental bodies formulate and implement policies to regulate and address societal and economic needs to improve health and prosperity. They must regularly assess these policies to ensure they remain effective and equitable while identifying areas for improvement.

Metrics are designed to objectively assess, report, and track policy performance against set targets or benchmarks. The quantitative insights they provide can help policymakers and stakeholders assess a policy’s impact and identify areas that require improvement, ultimately leading to better outcomes for citizens.^1^

While there is no globally uniform definition of a rare disease, in the United Kingdom (UK), a rare disease is defined as a condition that affects fewer than one in 2,000 people, i.e., fewer than 0.05% of the population. Given that over 7,000 rare diseases exist, more than 3.5 million people in the UK live with one or more rare diseases.^2^ Due to their rarity, limited research and expertise may exist on an individual condition, final diagnoses are often delayed, and care for people living with rare disease is frequently fragmented and inconsistent.^3^

To improve the lives of people living with rare disease, the Department of Health and Social Care (DHSC) published its first UK Rare Diseases Strategy in 2013. The aim was “to ensure that people living with rare disease have the best evidence-based care and treatment that our health and social care systems, working with charities and other organisations, our researchers and industry can provide”.^3^ In 2021, the UK Rare Diseases Framework superseded the original strategy. This new Framework outlined four national policy priorities for the next five years: (i) helping patients get a final diagnosis faster, (ii) increasing awareness of rare diseases among health professionals, (iii) better coordination of care, and (iv) improving access to specialist care, treatment and drugs.^2^ In England, the UK Rare Diseases Framework is implemented through the England Rare Diseases Action Plans,^4–6^ which DHSC reviews annually. Each Action is aligned with one of the four Priority Areas of the Framework. Evaluation is essential to assess whether and to what extent the England Rare Diseases Action Plans improve the lives of people with rare disease in England over five years (2021-2026). As implementation progresses, it is crucial to identify and agree on high-level metrics that policymakers and stakeholders can now use to monitor progress effectively.

This protocol outlines the design of an eDelphi study to develop a consensus on the most relevant and appropriate metrics for this purpose. The study will identify potential metrics and explore variations in consensus and priority levels across different policy domains and stakeholder groups.

This eDelphi study is part of a more extensive research programme funded by the National Institute for Health and Care Research (RareCare: www.rarecare.online). RareCare consists of two research strands: compiling high-level metrics for a portfolio-level evaluation of the England Rare Diseases Action Plans (Strand 1) and measuring time to diagnosis for people with rare diseases (Strand 2). The eDelphi study is part of Strand 1. Consilium Scientific (LO) and Coventry University (PAW) co-lead this Strand.

The Delphi method is a well-established and structured approach for developing consensus on issues that depend on expert judgment.^7^ This method is also particularly suitable for identifying key metrics for future use in a complex area such as rare diseases policy. The study’s findings are expected to provide evidence-based recommendations to policymakers, offering a robust set of high-level metrics for assessing the effectiveness and success of the England Rare Diseases Action Plans.

## 2. METHODS AND ANALYSIS

We will conduct a modified two-round eDelphi study to identify and build consensus on key metrics for assessing the effectiveness of the England Rare Diseases Action Plans.

The Delphi method was chosen because participants’ identities remain anonymous. This facilitates the free sharing of opinions and ensures that all judgments and perspectives are equally considered in the consensus process.

The eDelphi panel will assess candidate metrics for their relevance to the Actions, prioritise which ones will be most relevant as future high-level policy metrics, and consider their validity, reliability, interpretability, and the feasibility of data collection.

### 2.1 eDelphi Project Management Team

The eDelphi Project Management Team includes an epidemiologist (PAW), a health technology assessment researcher (AS), health policy and economic researchers (LO, JL, SL), two experts in rare disease policy, patient advocacy, and facilitating access to experts in the field (LB, JG), two clinical academics with recognised expertise in rare disease care (RL, PG), an academic ethics fellow (MS), a health psychologist with expertise in qualitative research (AWC), a design manager with expertise in methods to fully involve people in the design and development of services (NH) and a project manager with expertise as a global health advisor (ACS). PAW, AS, and NH have experience conducting and analysing Delphi studies.

Coventry University (CU) will lead the study, conduct, and data analysis. The eDelphi Project Management Team will review, interpret, and summarise the responses to each round, prepare feedback from the first round for the participants to consider in the second round, and attend a final meeting after round two during which any areas that would benefit from further exploration through qualitative research, including interviews, focus groups, or surveys are identified.

### 2.2 Patient and Public Involvement and Engagement

The UK Standards for Public Involvement will inform the study and incorporate lessons from the INSPIRE project.^8^ It will be guided by the principle of ‘no decision about me without me’ and, where possible, we will follow the National Institute for Health and Care Research (NIHR) INCLUDE roadmap,^9^ the INCLUDE Ethnicity Framework,^10^ ^11^ and the INCLUDE Socioeconomic Disadvantage Framework^12^ to emphasise the inclusion of underserved groups. We aim to adhere to the latest NIHR guidance on inclusive research, issued on 27^th^ November 2024.^13^

As mentioned, the eDelphi study is embedded in a larger project (RareCare: www.rarecare.online). RareCare has a Patient and Public Involvement and Engagement (PPIE) Group, chaired by LB of Realise Advocacy (RA). It consists of 11 members representing seven umbrella patient advocacy groups: CamRARE, Beacon, Metabolic Support UK (MSUK), Genetic Alliance (GA)’s SWAN UK, RARE Revolution, RareQoL, and Breaking Down Barriers (BDB). Together, these organisations support individuals with a broad spectrum of rare diseases, including inherited metabolic disorders, genetic conditions, and undiagnosed syndromes. The PPIE Group also includes organisations focusing on patients and families from underserved, diverse, and grassroots communities. Additionally, the group includes a representative from a network of healthcare professionals (Medics 4 Rare Diseases, M4RD).

The PPIE group co-produced the recruitment strategies and participant-facing materials and commented on the protocol.

### 2.3 Research Advisory Group

RareCare also has a Research Advisory Group (RAG), comprising 13 clinical and non-clinical members and two co-chairs (JG, MS). Members provide expertise and advice across clinical genetics, paediatrics, metabolic diseases, psychology, general practice, market access, healthcare commissioning, patient involvement in research, ethics, diversity and inclusion in research, disease policy, access and lived experience of rare disease. The RAG has reviewed and commented on the study protocol.

### 2.4 Choosing candidate metrics for the first round of the eDelphi study

The design of the eDelphi study commenced with a review of existing literature and available information to refine its scope, provide structure, minimise participant information overload, and improve the efficiency of the eDelphi study conduct. This included examining the monitoring and evaluation criteria specified in the England Rare Diseases Action Plans and evidence from the literature.

#### 2.4.1 Short-listing of current candidate metrics

Response rates of Delphi studies can decrease by 1.4 % (95% confidence interval: 0.3-2.5%) for every additional ten items included.^14^

To balance comprehensiveness with maintaining response rates and minimising participant fatigue, we first extracted all action-specific monitoring and evaluation criteria from all England Rare Diseases Action Plans published in 2022, 2023, and 2024,^4–6^ and reviewed and refined them.

Five reviewers (JG, PAW, MS, PG, RL) then independently assessed each criterion’s suitability as a metric, using a scoring system of 2 (‘Keep’), 1 (‘Change’), or 0 (‘Remove’). Average scores were categorised into five levels: unacceptable (0– 1), poor (1.01–2), acceptable (2.01–2.99), good (3–3.50), and very good (3.51–4). Reviewers also considered each metric’s potential impact on health inequalities and identified any Actions for which additional metrics might be assessed.

The eDelphi Project Management Team considered this information in three online workshops to decide which of the original 88 short-listed metrics should advance to the first round of the eDelphi study. During these sessions, the research team also considered whether each metric was clearly worded, transparent, and capable of reflecting change over time.

#### 2.4.2 Evidence Review

In parallel, three researchers (JL, SL, and FM) conducted a scoping review of peer-reviewed and grey literature (e.g., documents, videos, lectures, and seminars) to identify additional candidate metrics. This evidence review focused on the four Priority Areas identified in the 2021 UK Rare Diseases Framework.^2^ A separate search was conducted for each area. Databases searched included PubMed, Google Scholar, and SciSpace (an AI-based literature search portal).

Inclusion criteria were that a resource should (i) specifically address rare diseases, (ii) align with one of the Priority Areas, and (iii) have been published since 2014. Non-English language resources were considered only if published in Spanish or Italian (spoken by the team) and if they provided findings that applied to the UK healthcare system. Extracted candidate metrics were reviewed by at least one other team member, with disagreements resolved through discussion.

**Figure 1.**
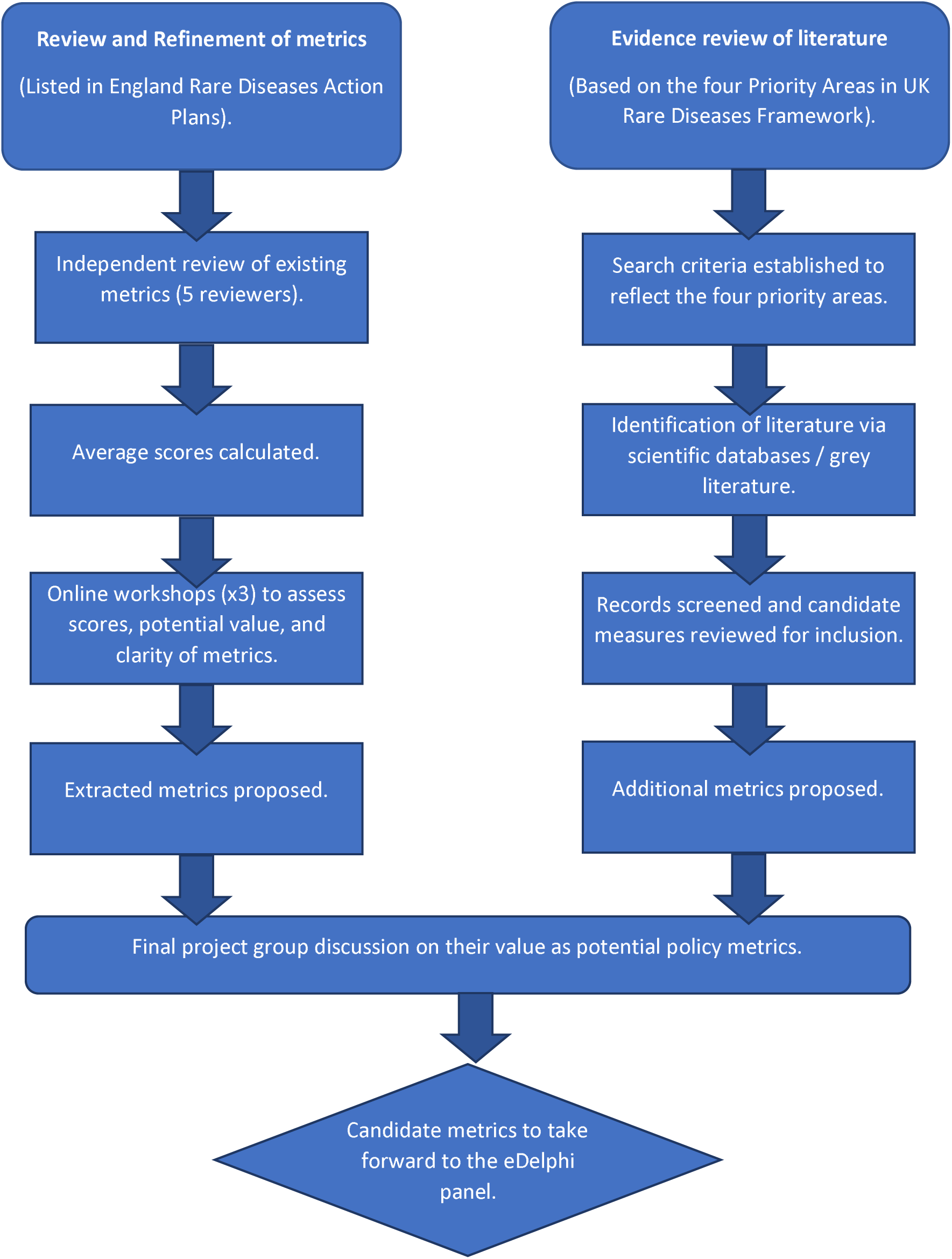
Flowchart detailing decision-making for candidate metrics.

#### 2.4.3 Compiling the final list of candidate metrics for the eDelphi panel

Of the 89 initial candidate metrics extracted from the Action Plans, 55 were excluded because they either did not reflect change over time or corresponded to a completed action that was no longer relevant. The remaining 34 metrics were produced by rephrasing or dividing existing measures to address gaps or improve clarity. Another 18 metrics were added from the evidence review. A further 12 were added and three were removed by expert consultation. The final 61 candidate metrics will be presented to the eDelphi panel in Round 1.

### 2.5 Design of the study materials

All participant-facing materials are written in plain English and pretested to ensure they are clear, understandable, and appropriate for the target population. They are tailored to the specific needs of each stakeholder group to ensure they are as relevant and accessible as possible.^15^ The PPIE Group reviewed the materials for the panel members with lived experience of rare disease, and their feedback has been incorporated to ensure comprehension and flow. In addition, the questionnaires have been piloted to assess comprehension and time to completion further, and adjustments have been made accordingly.

The eDelphi study materials, including the participant information sheet and the consent form, will be hosted on the Qualtrics (Qualtrics, Provo, UT, USA) platform, which is GDPR compliant and certified to the ISO 27001 standard.

#### 2.5.1 Participation information sheet

The Participation Information Sheet outlines the research’s purpose and details the organiser and funder. It explains that participation is voluntary, and invitees are not obligated to participate. The sheet also describes what will happen if someone chooses to participate, why the individual has been invited, the benefits and potential risks, the type of information being collected, and the lawful basis for processing it. It further explains what will happen to the research results, who will have access to the data, where it will be stored, that only pseudonymised data will be shared, and what the eDelphi study entails. Contact details for the researcher and a point of contact for any questions or concerns are also included.

#### 2.5.2 Informed consent form

The informed consent form will ask participants to confirm that they:

- Have read and understood the Participant Information Sheet and had the opportunity to ask questions.
- Understand that their responses relating to rare diseases must be based on experiences from living in England.
- Are informed about what will happen to their data after the research project concludes.
- Understand that participation is voluntary and can be withdrawn at any time before data analysis without providing a reason.
- Are aware that the research results will be used in academic reports and publications.
- Understand that their participation and data will be anonymised unless they explicitly consent to their name being acknowledged in the final scientific publications.
- Do not have a financial conflict of interest and have considered whether any personal or professional interests in how the impact of the UK Rare Diseases Framework is measured could influence their objectivity and ensure that their responses remain impartial.

Participants will then be asked to confirm their consent and provide their contact details. At the end of the first-round questionnaire, participants will be asked to confirm that they agree to participate in the second round of the eDelphi study.

#### 2.5.3 Demographic questions

Selected demographic questions will collect information about each participant, including their age group, gender, ethnicity, geographical region, and the stakeholder group(s) with which they identify. Participants who indicate they are part of the lived-experience group will also be asked whether the rare disease is genetic/non-genetic, what its main health consequences are, and whether it is stable, progressive, or life-limiting. For participants who indicate that they are professionals working in rare diseases, additional questions will request information regarding their main field of work/expertise, how many years of experience they have working in rare diseases, and any experience and/or involvement in assessing or developing rare disease policies/guidelines.

#### 2.5.4 Round 1 questionnaire

The questionnaire will begin with a brief overview of the UK Rare Diseases Framework and will be organised into sections covering the four Priority Areas. Each section will introduce the Priority Area. This will be followed by questions about the associated Action Plans and their short-listed metrics. For example, in terms of ‘Better coordination of care’, one set of candidate metrics might relate to the ‘virtual consultation toolkit to help healthcare professionals in planning and delivering online consultations’. The panel members will be asked to rate the importance of the candidate metrics extracted to date on a Likert scale ranging from 1 to 9, with the option of declaring “Unable to rate”.^15^ ^16^ Ratings of 1 to 3 will indicate that the metric is “not valuable,” 4 to 6 will indicate “valuable but not critical,” and 7 to 9 will indicate that it is “extremely valuable” to assessing progress. Three optional open-ended questions will allow the panel members to suggest any rewording of the metrics, explain any of their answers, and suggest any additional measures that may help to measure progress in the priority area. At the end of the questionnaire, the panel members will have a final opportunity to propose further high-level policy metrics for assessing the England Rare Diseases Action Plans. While the rating questions will be mandatory, all open-ended questions will remain optional to encourage participation without imposing an undue burden.

#### 2.5.5 Round 2 questionnaire

The questionnaire for the second round of the eDelphi study will be based on analysis of the first-round responses. It will follow a similar structure to the first-round questionnaire but will focus specifically on candidate metrics for which consensus on inclusion or exclusion was not reached. Additionally, it will include any new metrics suggested by the panel experts during the first round. Metrics that received a consensus of low value will be excluded unless the panel members indicate that the wording requires significant clarification. For other candidate metrics, each panel member will be presented with their rating from Round 1, alongside the median rating from other members within their stakeholder group, the overall panel rating, the interquartile range, and a summary of all participant comments.^17^

Presenting information in this way will allow each panel member to see how their responses have contributed to reaching the predefined consensus level. This will encourage them to reflect on their earlier answers and decide whether to revise their judgments.^17^ Participants can also provide a rationale for their ratings, suggest rewording, or offer additional comments.

The identity of individual participants and their responses or comments will remain confidential throughout the study. However, participants may choose to be acknowledged for their contributions to the study outputs once the study has concluded. They will also be asked whether they can be recontacted for a summary of the results.

### 2.5 Sampling frame

We have defined a purposive sampling frame to recruit a diverse cross-section of 50 individuals with lived experience of rare diseases and 50 professional stakeholders in the field. This sample size allows for 20% attrition in Round 2 of the eDelphi study. By including people with lived experience as half of the panel, we will increase the likelihood of identifying metrics that genuinely reflect the policy’s impact on the lives of people living with rare disease.^18^ Because the 7,000 rare diseases fall under a range of medical specialities and vary widely in age of onset, symptoms, severity, and the populations affected, individual Rare Disease Action Plans may affect individual panel members differently or not at all. Therefore, we have also tailored the sampling frame to ensure that the final selection of high-level metrics presented to the eDelphi panel members will be as relevant as possible to the broader rare disease community. Broad panel membership should enhance the credibility and acceptability of the metrics finally identified for assessing government policy in this field,^19^, although the heterogeneity of the sample may limit clear consensus in some areas.^20^ The people with lived experience of rare disease or their carers and the professional stakeholders recruited may exhibit bimodal response patterns in some areas, and the reasons for this heterogeneity will be explored.

#### 2.5.1 Recruitment of people with lived experience of a rare condition

People with lived experience as either living with a rare disease or being a carer will be recruited via the PPIE Group. First, because of the wide variety of rare diseases and their symptoms, a targeted approach will be adopted to ensure access to individuals with diverse lived experiences, covering various conditions and stages of the patient journey. Second, although laypeople might feel as though they have less of a direct stake in the policy framework than professional stakeholders, they are less likely to be biased in their opinions.^21^ Therefore, there are clear benefits of utilising a confidential recruitment process to access these groups’ ‘real’ views, unaffected by ‘expert’ opinion.^21^ Finally, there is evidence that visible socio- demographic status and other contextual factors are more likely to influence a layperson’s rating compared to those of experts. This will support a strategy of separate feedback at stage 2.

People with rare disease and their carers will be recruited through patient advocacy groups with diverse remits, including Beacon, Breaking Down Barriers (BDB), CamRARE, SWAN UK (part of Genetic Alliance), Metabolic Support UK (MSUK), and RARE Revolution. Despite such organisations typically drawing from a relatively limited spectrum of patients, this recruitment strategy should provide a less biased sample than recruitment via specific clinical routes and include more informed, proactive, and engaged individuals.^22^ The aim is to recruit six young people (16–25 years old), six adults with a rare disease that had a paediatric onset, six adults with a rare disease that manifested in adulthood, six older adults (60 years and older), eight carers of adults with a rare disease, and 18 carers of children with a rare disease.

Overall, the aim is for 75% of participants to have or care for someone with a genetic condition and 25% to have or care for someone with a non-genetic condition. Special attention will be paid to recruiting underserved groups, following the INCLUDE^8^ and INVOLVE^9^ ^11^ ^12^ guidelines and frameworks. The final eDelphi panel will therefore include representation from people who are: (i) carers according to disease condition (stable, progressive or life-limiting); (ii) live with different types of conditions (invisible conditions, intellectual disability, visual difference, sensory impairment, physical disability, or conditions of sex development; and (iii) live in geographical regions (London, South West, East Midlands, West Midlands, North West, North East). We will emphasise the importance of obtaining a diverse sample of people living with rare diseases/carers within each patient advocacy group. For the same reason, we will monitor the overall recruitment patterns and aim to adjust where beneficial.

Participants will receive a £25 shopping voucher per questionnaire to compensate them for their time and effort in the two-round eDelphi study.

#### 2.5.2 Recruitment of professionals with relevant expertise or experience in rare diseases

CS will coordinate the recruitment of professional stakeholders, with support from the PPIE Group, RAG, and RA, to identify experts for the eDelphi panel.

Professional stakeholders will be purposively selected based on their expertise and knowledge in healthcare policy, the rare disease sector, or relevant aspects of the healthcare system. These stakeholders will include representatives from NHS management (national and regional commissioning), healthcare professionals, industry, education, patient advocacy groups, the broader health and care system, and the data and research sectors. A sampling frame showing targets for separate groups will be developed in advance. Efforts will be made to ensure diversity in the geographical locations of stakeholders.

Identifying potential eDelphi panel members will involve reviewing scientific and medical literature, professional networks, presenters at rare disease conferences and events, those involved in Framework development, delivery partners, and end users of the policy metrics (e.g., the Department of Health and Social Care).

As a gatekeeper, CS will send invitations and directly monitor target numbers. Members of the RareCare team, RAG, and PPIE Group will not be eligible to join the eDelphi panel, ensuring the findings remain free from the potential influence of pre-existing opinions or competing interests.^23^

#### 2.5.3 Eligibility criteria

To be eligible for participation, individuals must:

– Live in England (if they are part of the ‘lived experience’ group, i.e., those living with rare disease and their carers).
– Have experience with rare diseases through work relevant to England (for professional stakeholders).
– Be 16 years or older.
– Have no self-declared financial conflicts of interest in assessing the impact of the England Rare Diseases Action Plans
– Not be a member of the RareCare team, RAG or PPIE Group.
– Be able to read and write in English.
– Have access to the internet and a device to complete the questionnaire.

### 2.6 Administration of the questionnaires

The gatekeepers (seven patient advocacy group members of the PPIE Group and CS) will email selected experts and invite them to join the eDelphi panel between March and May 2025. The invitation will contain a brief overview of the study and request an initial response if interested in contributing. Those who report an interest will be added to the global target number, and the gatekeeper will send a link to the participant information sheet and an informed consent form hosted on the Qualtrics platform. For people with lived experience of rare disease, if the recruitment target is not met, the patient advocacy groups will be informed by RA and asked to invite additional individuals. For professional stakeholders (i.e., researchers, healthcare professionals, policymakers, commissioners, delivery partners, and industry representatives), we will request that any individual who declines participation suggest alternative experts to approach.

Once a participant provides their content, they can access the questionnaire. Each round will be open for at least two weeks, and if response rates are low, this will be extended by a week.^24^ Up to three reminders will be sent to participants who have not completed the questionnaire.

We estimate that data analysis for Round 1 and preparation of feedback and questions for the next round (including possible rephrasing or refining of the candidate metrics and adding those newly suggested) will take three weeks. Once completed, the second-round questionnaires will be made available to those participants who consented to the second round.

### 2.7 Data analysis

#### 2.7.1 Analysis of consensus and responses

Consensus will be based on the levels of participant agreement and disagreement. Following the COMET (Core Outcome Measures in Effectiveness Trials) Initiative,^15^ consensus for inclusion will be achieved if 70% or more participants in both groups (people with lived experience and professionals) rate a metric as ‘extremely valuable’ (i.e., 7, 8, or 9 on the Likert scale) and fewer than 15% rate it as ‘not valuable’ (i.e., 1, 2, or 3 on the scale). Conversely, consensus for exclusion will be reached if 70% or more rate the metric as ‘not valuable’ and fewer than 15% rate it as ‘extremely valuable.’ A candidate metric will be accepted for inclusion or exclusion in the final selection if both groups reach a consensus in the same direction. If either group fails to meet the consensus criteria, the metric will remain under consideration.

The eDelphi Project Management Team will meet between Rounds 1 and 2 to assess whether any suggested new metrics fall within the study’s remit and would be suitable to measure progress mid-point for policymakers and other stakeholders. If deemed appropriate, their phrasing will be finalised for Round 2.

Round 2 data will be analysed as above. In addition, the stability of answers across the rounds will be assessed using descriptive analyses, such as the spread and measures of central tendency across the responses. Their determinants will be assessed using stratified analyses, including by stakeholder group.

#### 2.7.2 Analysis of open-text comments and the suggested new metrics

The rationale provided in response to open-ended questions following each candidate metric, any descriptions of suggested new metrics, and final summary free-text comments will be analysed using thematic content analysis.^25^ This approach focuses on identifying text-based meanings and patterns within the ‘thematic units’.^26^

A hybrid approach to analysis will be used. An initial deductive approach will use the four priority areas specified in the England Rare Diseases Framework^2^ to code texts related to the short-listed metrics. A codebook to classify data into predefined categories will be iteratively developed. These will be applied to the coded data using NVivo software (Lumivero, Burlington, Massachusetts, USA) to assist in identifying quantitative patterns and themes. Inter-coder reliability will be established via collaborative coding sessions with a second researcher (AWC, NH).

Following Round 1, the eDelphi Project Management Group will also review the rationales derived from the analysis of open-text comments. Final summaries of these, along with retained and new metrics, will be agreed upon for inclusion in Round 2.

#### 2.7.3 Final review of eDelphi study findings

Following the Round 2 data analysis, all policy metrics demonstrating a clear consensus will be identified.

Subsequently, a research plan for qualitative data collection—using methods such as semi-structured interviews, focus groups, or surveys—will be developed. For any Action Plan where consensus on none of the metrics has been reached, further exploration will be conducted through qualitative studies involving individuals with lived experiences and relevant stakeholders. Candidate metrics with mixed ratings may also be examined using this approach.

The analysis of participants’ rationales for rating the candidate metrics, as captured during Round 2, will inform the interview or focus group schedule design. A separate protocol for this qualitative phase will be prepared and submitted for review by the ethics committee.

## 3. ETHICS AND DISSEMINATION

### 3.1. Data handling and confidentiality

Participant confidentiality and all data will be handled following CU Policies, Standards, and Procedures, including its Research Ethics and Integrity Statements and Research Privacy guidelines (available at https://www.coventry.ac.uk/gdpr-and-dataprotection/privacy-notices/). These comply with the General Data Protection Regulation 2018 and the Data Protection Act 2018.

Participant data will be collected through Qualtrics and stored as raw data on CU’s Microsoft Teams for Research site. The data will be stored in separate, password-protected, and encrypted files: i) questionnaire data and ii) personal data.

Identifiable participant information will be accessible only to two CU staff members (AWC, NH), who will assign a unique participant identifier number to each participant. Other CU staff (PAW, AS) can only access pseudonymised data, including participant identifier numbers. The pseudonymised data will be imported into statistical software (e.g., Stata or SPSS) for quantitative analysis or into Word or NVivo for qualitative analysis by the CU team.

The raw dataset will remain within CU and not be shared externally, except for audit purposes, if required. Only authorised CU staff will access the data, ensuring confidentiality and security. Non-identifiable summary data (e.g., newly suggested metrics, consensus decisions) will be shared with other project team members, the RAG, and the PPIE Group and included in any publication. To maintain anonymity, any group presented separately will consist of at least five individuals with similar characteristics.

A dedicated project space on Microsoft Teams for Research will be used for secure storage and data sharing with project colleagues. This managed storage, provided by CU Digital Services, has scheduled backups. All project data will be stored following CU policy, in password-protected files within Microsoft Teams for Research. CU is certified for Information Security ISO 27001 Standard, Cyber Essentials and Cyber Essentials Plus.

### 3.2 Managing risks to panel members

PAW, AWC, and NH, who are responsible for the study conduct and analysis, have undertaken Good Clinical Practice and Equality, Diversity and Inclusion training.

Although participants’ distress levels are likely to be low in this study, given that they are not being asked to offer personal accounts of their experiences in the rare disease world, the subject matter remains a sensitive and emotive topic for some individuals. The PPIE Group and RAG have advised on the appropriateness of the research for the target audience and minimisation of distress, e.g., via the wording in the participant information sheets and questionnaires.

The participants will receive a list of resources (e.g., helplines, counselling, and support services) tailored to the rare disease community.

The participant information sheet will include researcher contact information and a clickable link to email addresses to make it easy for participants to contact the research team if they have concerns or need additional support. If a participant contacts the researchers directly, demonstrating distress, the researcher will follow the steps outlined in the distress protocol flow chart. The gatekeepers will be advised to copy parents, carers or guardians when inviting a 16 or 17-year-old for participation.

The participants will be assured they can withdraw their personal data without consequences for their care, treatment, or job role. They will also be assured that they can withdraw their answers to the Round 1 questionnaire until Round 2 begins; and their Round 2 data until the data analysis is complete.

### 3.3 Ethics approval

The Coventry University Group Research Ethics Committee has approved the study on 19 March 2025 (Project code: P183377). The questionnaires will only be distributed to participants who have provided informed consent.

### 3.4 Dissemination plan

The findings will be disseminated through conference presentations, policy briefs for government agencies and policymakers, and an open-access, peer-reviewed journal.

Reporting will follow the ACCORD (ACcurate COnsensus Reporting Document) guidance.^27^ A plain-language summary of the results will be available on the RareCare website, and it will also be shared with gatekeepers and study participants who opt in to receive it.

Given the small field of rare diseases, we will report findings for larger stakeholder groups (minimum of five people) to minimise the risk of indirect identification.

Anonymised group-level data will be stored in an open-access repository and assigned a DOI to ensure long-term accessibility and ease of citation.

## Data Availability

Given the small field of rare diseases, we will report findings for larger stakeholder groups (minimum of five people) to minimise the risk of indirect identification.
Anonymised group-level data will be stored in an open-access repository and assigned a DOI to ensure long-term accessibility and ease of citation.

## AUTHORS’ CONTRIBUTIONS

PAW, AS, LB, JG, and AWC conceptualised the study and designed the research. LO, JG, and PAW secured the funding. PAW, AS, ACS, SL, and JL conducted the literature review. JG, LB, MS, PG, and RL provided rare care expertise. JG and MS coordinated input from the RAG, LB from the PPIE group, and JG and LO from professional stakeholders. PAW, AS, JG, LB, MS, and RL critically reviewed the extracted candidate metrics. SL, JL, ACS, AWC, and LO synthesised the metrics review data. PAW, AS, LB, AWC, and ACS designed the data collection and management procedures. PAW, AS, and AWC planned the data analysis. PAW and AS drafted the manuscript. All authors critically reviewed the manuscript for important intellectual content. All authors read and approved the final manuscript and agreed to be accountable for all aspects of the work. PAW is the guarantor.

## COMPETING INTERESTS STATEMENT

All authors have completed the ICMJE Uniform Disclosure Form. They declare funding from the National Institute for Health and Care Research and no support from any commercial organisation for the submitted work. Coventry University has provided in-kind support for PAW and AS in relation to their contributions to this study. JG and LB, through Realise Advocacy Ltd, have received financial support from patient advocacy groups and industry stakeholders for consultancy services related to patient involvement in drug development and Health Technology Assessment. JG leads JG Zebra Consulting, which provides international and national strategic consultancy services specialising in rare diseases and novel therapies, focusing on market access, public policy, and stakeholder engagement. LB chairs the Board of Trustees at Medics 4 Rare Diseases. LB and RL are members of the Rare Diseases Advisory Group (RDAG), which advises NHS England and the devolved administrations on the development, implementation, and national commissioning of highly specialised services for rare diseases. PG is a co-inventor on five filed patents in rare disease therapeutics and has received support for meeting attendance and medical writing.

## FUNDING STATEMENT

This work is independent research funded by the National Institute for Health and Care Research (NIHR) [RareCare, NIHR205983]. The views expressed in this publication are those of the author(s) and not necessarily those of NIHR or The Department of Health and Social Care.

## ACKNOWLEDGEMENTS

We thank Nikki Holliday (NH) for her advice on data collection and management procedures and Federica Mirto (FM) for contributing to the literature review. We are also grateful to the PPIE Group and RAG for their feedback on the protocol and active contributions to the patient-facing materials. Additionally, we thank Professor Stephen A. Morris for his insights on the ‘Coordination of Care’ priority area of the England Rare Diseases Action Plans.

## ORCHID IDs

Professor Petra A Wark

Dr Leeza Osipenko

Josie Godfrey

Lindsay Birrell

Dr Aimee Walker-Clarke

Dr Andrea Caputo Svensson

Scott Lewis

Jack Lewis

Dr Mark Sheehan

Dr Robin H Lachmann

Professor Paul Gissen

Professor Ala Szczepura

## Notes

### Clinical Protocols

https://www.isrctn.com/ISRCTN41639707

### Author Declarations

The Coventry University Group Research Ethics Committee has approved the study (P183377).

